# Characterising People who inject drugs, and association with HIV infection: A Situation Analysis in Kampala city, Uganda

**DOI:** 10.1101/2024.05.31.24308282

**Authors:** Peter Mudiope, Bradley Mathers, Joanita Nangendo, Mutyaba Samuel, Byamah Brian Mutamba, Stella Alamo, Nicholus Nanyenya, Fredrick Makumbi, Miriam Laker-Oketta, Rhoda Wanyenze

**Author notes:** Peter Mudiope: **Corresponding Author, Email of Corresponding Author:**. **Authors’ contribution:** PM, FM RW, ML, JN, and MB wrote the study concept and design. PM, MS, and JN carried out data collection. PM, MS, and NN performed statistical analyses and interpretation of the data. All authors participated in writing the manuscript and read and approved the final manuscript.

## Abstract

**Background:** Uganda has implemented targeted interventions to address the rising burden of injection drug use, yet barriers persist in reaching persons who inject drugs (PWID). This study describes the characteristics of people who inject drugs, physical and mental health states, and associated risk behaviors, to inform the designing of programs that are tailored to client’s needs and preferences.

**Methods:** A cross-sectional survey was conducted between August and December 2023 at selected hotspots in Kampala, interviewing 499 PWID aged ≥18 years. Data was collected using a semi-structured questionnaire administered by peer educators and Uganda Harm Reduction Network (UHRN) counselors. Measurements included socio-demographics, injecting drug use and sexual risk behaviors, and medical history. HIV serostatus was dtermined by self-report and testing for participants who had no recent history of testing and consented to be tested. Binary logistic regression was used to establish the relationship between HIV infection and risky drug- and sexual behaviors of PWID.

**Results:** Participants were predominantly Ugandan (95.2%), male (73.2%), unmarried (55.9%), unemployed (81.8%), with higher levels of education and varying ages. Mental disorders were prevalent, with 48.7% reporting at least one underlying condition, including depression (30.8%) and anxiety (9.6%). Physical health issues were also noted, with reported cases of fever (32.9%), cough (32.5%), malaria (22%) and sexually transmitted infections (STIs) (15%).

Regarding drug use patterns, the majority (82.6%) were introduced to drugs by close acquaintances, with 70.9% categorized as people who inject drugs. HIV prevalence among injecting drug users was 3.7%, with higher rates among females (8.4%) and non-Ugandans (16.7%). Being female and experiencing difficulty accessing sterile injection materials were associated with HIV-positive status, highlighting the complex interplay between socio-demographic factors, risk behaviors, and HIV infection among individuals with injecting drug use Disorder in Uganda.

**Conclusion:** Our study provides a comprehensive insight into the socio-demographic, mental, physical health, and HIV risk behavoir of PWID in Kampala, Uganda. The findings indicate significant vulnerabilities to injecting drug use, mental disorders, and high-risk behaviors that predispose this population to HIV infection. Despite a low HIV prevalence compared to previous estimates, the interplay between drug use, risky injecting practices, and sexual behaviors suggests an urgent need for targeted interventions to address these intertwined challenges.

## Introduction

Recent global data indicate an increasing supply and demand for illicit drugs. The United Nations Office for Drugs and Crime (UNODC) reports that ilicit drug use increased by 23% from 240 million in 2011 to 296 million in 2021, of whom 13.2 million people were injecting drugs (1). Louisa Degenhardt et al (2023) estimated that 14·8 million people are drug injectors in the 190 countries that contribute more than 99% of the global population(2). The rising burden of drug use including injecting, continues to harm the economic, health, and social sectors(1). The high prevalence of HIV and Hepatitis B and C infections in Sub-Saharan Africa has in part been attributed to the observed increase in injection drug use (3–5). This problem is compounded by a high occurrence of infectious diseases, as well as a myriad of socio-demographic, mental health, and other general physical health issues among PWID. Studies have reported injection drug users to be mainly men, not married, unemployed, often young with varied education levels, and prone to high risk of sexual and social crime (3). Concurrent mental disorders with OUD also present a dual burden for PWID. A high prevalence of depression and anxiety among people with OUD was reported in sub-Saharan Africa (6). However the prevalence of mental disorders among PWID varies widely based on the method of ascertainment(7–9). A recent study indicated that the prevalence of depressive disorders varied from as low as 3·44% to a high 66% in the general population (2).

In Uganda, recent estimates put the number of injecting drug users at 7,356 with an HIV prevalence of 16% compared to 5.6% in the general adult population, making it the second highest contributor to new infections among key populations. and is the second largest contributor to new infections among key populations in Uganda (10). As Uganda strives to end AIDS by 2030, critical attention is needed to reduce HIV transmission among PWID and other key populations. Acknowledging the burden of HIV among PWID, Uganda adopted the WHO/UNODC recommended HIV prevention interventions for PWID as outlined in the national strategic plan(11, 12). Working with community partners support led by the Uganda Harm Reduction Network (UHRN), the Global Fund and other UN health development partners supported a project that distributed clean needles and syringes in Kampala and Mbale cities which account for more than half of the population of injecting drug use in Uganda (13). In 2020, with PEPFAR support, Uganda set up the first MOUD clinic at a centralized clinic located at the psychiatric hospital in Kampala capital city. Hitherto another clinic, has been established in Mbale city in the Eastern part of the country. Despite these efforts, the scope of service and geographical reach of HIV prevention, care, and treatment services for PWID is far from realizing the targeted 90%. The barriers to optimally reaching PWID with such services have previously been described to act at multiple levels including social, physical, economic, or political (14, 15). Key barriers include; stigma, peer influence, criminalizing of drug use, lack of transport to the service points, comorbidities such as mental disorders, limited knowledge of available services, and limited capacity to provide services among others(14, 15).

The one-size-fits-all services delivery approach has proved ineffective, as challenges in enrolling and retaining clients in harm reduction programs persists. There is a need to improve service delivery by offering differentiated harm reduction services based on client needs and preferences. Such program shifts require understanding the risk profiles of PWID regarding their socio-demographics, drug use, risk behavior, and criminal and mental health among others. PWID risk profiles are heterogeneous and given the limited resources, tailored services based on PWID needs and preferences are required (16). In Uganda, evidence on the relationship between PWID’ characteristics and their risk of HIV infection is still limited. Studies have been primarily exploratory with small sample sizes and have not fully captured the variations in participants’ characteristics related to their risk of HIV infection and access to HIV prevention services (17). Knowledge of the varying profiles of PWID is crucial for the implementation and uptake of risk reduction interventions. This study aimed to describe the characteristics of people who inject drugs, physical and mental health, and HIV risk behaviors to inform the design of programs that are tailored to client’s needs and preferences.

## Methods

### Study design & setting

This cross-sectional survey was implemented from August 2023 to January 2024 at selected hotspots in Kampala City. The study was conducted with the active involvement of the Uganda Harm Reduction Network(UHRN), a PWID community-based organization that provides harm reduction services to PWID at different drop-in centers and hotspots in Kampala.

#### Study population, sampling and sample size

In total, 38 hotspots frequented by injecting drug users were conveniently selected based on the safety, security status, and willingness of local leaders to permit data collection. We recruited 499 injecting drug users aged 18 years and above who had a history of at least one drug injection in the previous three months, as established by history and the manifestation of recent skin injection marks. Recruitment within the community utilized peer educators who mobilized participants at hotspots or DICs to participate in the study. Interviewed participants were also encouraged to refer other users within their network, helping the study to achieve an adequate sample size. All the 499 injecting drug users were interviewed after providing informed consent.

#### Study procedure

The data were collected using a semi-structured questionnaire and keyed directly into open data kit (ODK) database software(18). The questionnaire was piloted among PWID attending the MOUD clinic at Butabika National Referral Mental Hospital. The MOUD clinic staff were trained to collect data. Two additional peers were recruited and trained to mobilize venue owners and participants to support and participate in the study respectively. During visits to the hotspot or drop-in centers, the data collectors administered the questionnaire in the participant’s preferred language. The data was entered directly into the ODK, where the data manager provided the quality control feedback for redress by data collectors.

To increase the likelihood of participation, we leveraged existing social networks, encouraging initial participants who would be willing to participate in the study. We prioritized flexible scheduling of interviews to accommodate the participant’s availability while considering their mental status, work schedules, treatment appointments, and other commitments. Due to legal restrictions and the potential risk of arrest, accessing PWID posed significant challenges. Therefore, we prioritized the safety and well-being of both participants and data collectors. Throughout the data collection process, participants were assured of their safety and the confidentiality of their information. They were also informed about the importance of their participation in the study for informing public health interventions. Collaborating with UHRN staff, venue owners, and peer educators helped to reassure participants about the confidentiality of their involvement and alleviate concerns about potential legal repercussions.

#### Study measurements

Participants’ sociodemographic characteristics included age, sex assigned at birth, religion, nationality, education level, monthly income, and source. Regarding substance use, participants were asked which substances they often used/injected in the previous 30 days, the lifetime duration of use, reasons for use/injection, and the frequency of use/injection. Questions on the frequency and use of flash blood syringes, sharing needles/syringes, and frequency of experiencing overdose were asked to establish the injection risk behaviors. Sexual risk behaviors were assessed by asking about the sexual encounters (vaginal or anal) with non-regular partners and condom use in the last 30 days. Participants were considered to have consistently used condoms if they used condoms for ALL times during during all the sexual encounters. Sexual risk was also assessed by asking participants if they had sex with someone else in exchange for money, drugs, or any other commodity as the term for payment. Physical illness in the past three months and screening for HIV, TB, and hepatitis were also assessed. Those who reported being HIV positive and had their status confirmed from clinical records were not required to undergo additional testing. For others, HIV status was determined using the Determine™ HIV-1/2 rapid test, with confirmation via the Bioline HIV1/2 3.0 rapid test, following national guidelines (18). Mental illness was assessed through self-reporting, asking if particpants were or had been on treatment for a mental health condition. Sexual gender-based violence(SGBV) was evaluated by asking if a participant had ever been forced to have sexual intercourse (19). Other factors assessed included the history and frequency of incarceration.

#### Statistical Analysis

Baseline socio-demographic, mental, and physical health-related, drug use behaviors and sexual risk profiles were summarised using medians and interquartile ranges for numerical variables and proportions for categorical variables. Binary logistic regression, was used to determine the association between the independent variables (socio-demographic, drug use, and sexual-related risk profiles) and the HIV status of participants.

During bivariate analyses, variables that had p values <0.1 or known association with HIV serostatus, were selected for inclusion into multivariate analysis. In multivariate logistic analysis level, we adjusted for confounding using the backward stepwise method based on p-values, removing most non-significant variables and interaction terms from the model until only significant variables remained. The variables with p-values<0.05 were reported as significantly associated with HIV status. Adjusted Odds ratios (AORs) and the corresponding 95% confidence intervals (CI) were reported. All analyses were performed using the STATA version. 14.2 (College Station, Texas).

#### Ethical Considerations

This evaluation was approved by the Makerere University School of Public Health, the Higher Degrees Research and Ethics Committee, the Uganda National Council of Science and Technology (UNCST), and the administration of Butabika Hospital.

## Results

A total of 499 participants were interviewed, predominantly male (73.2%), of Ugandan nationality (95.2%), unmarried (55.9%), and not in regular employment (81.8%). The median age was 31 years, interquartile range (IQR), (26–36). A significant percentage of participants resided either alone (48.1%) or with a partner or friend (46.1%), identified with the Christian faith (63.5%), and had achieved at least a secondary level of education (60.3%). A considerable proportion (71.5%) reported a monthly income of between 50,000/= to 500,000/= Ugandan shillings inclusive, while the majority (72.3%) disclosed a history of prior detention or incarceration(**Table 1**).

**Table 1.** Socio-demographic characteristics of people who use illicit drugs in Kampala, Uganda (n = 499)

| Variable | Total (n= 499) |
| --- | --- |
| **Sex** |  |
| Male n (%) | 365 (73.2) |
| Female n (%) | 134 (26.8) |
| **Age -median (interquartile range-IQR)** | 31 (26 to 36) |
| 18-24yrs n (%) | 92 (18.4) |
| 25-29yrs n (%) | 137 (27.4) |
| 30-34yrs n (%) | 127 (25.5) |
| 35yrs &above | 143 (28.7) |
| **Highest education level attained(n=424)** |  |
| No education n (%) | 60 (12) |
| Primary level n (%) | 138 (27.7) |
| Secondary n (%) | 226 (45.3) |
| Post-secondary n (%) | 75 (15) |
| **Married Status** |  |
| Married | 26 (5.2) |
| Cohabiting | 59 (11.8) |
| Separated/Widowed | 135 (27.0) |
| Un-married | 279 (55.9) |
| **Religion** |  |
| Christian(Anglican/Born-again) | 196 (39.3) |
| Christian(Catholics) | 121 (24.2) |
| Islam | 174 (34.9) |
| No religion | 8 (1.6) |
| **Nationality** |  |
| Ugandan | 475 (95.2) |
| Non-Ugandan | 24 (4.8) |
| **Staying with** |  |
| Alone | 240 (48.1) |
| Parent/Children | 29 (5.8) |
| Partner/Friend/relative | 230 (46.1) |
| **Employment status(source of income)** |  |
| Regular employment | 91 (18.2) |
| Informal casual work | 272 (54.5) |
| Other activities(such as sex work) | 126 (25.3) |
| No source of income | 10 (2) |
| **Monthly Income(UGX)** |  |
| <=50,000/= | 117(23.5) |
| >50,000-500,000ugx | 303 (60.7) |
| >500,000/= | 79 (15.8) |
| **Ever been imprisoned for more than 24 hours in life** |  |
| Yes | 361 (72.3) |
| No | 135 (27.1) |
| Decline to respond | 3 (0.6) |

### Mental health illnesses

In total, 243 individuals (48.7%) disclosed having at least one underlying mental illness, among whom 55 (11%) were undergoing treatment, and 64(12.8%) reported more than one existing mental condition. Among those who reported mental illness, 48 (9.6%) acknowledged co-existing anxiety, with five of them receiving treatment; 154 (30.8%) indicated co-existing depression, with 39 (7.8%) undergoing treatment. Furthermore, out of the 33(6.6%) individuals who reported co-existing Attention Deficit Hyperactivity Disorder (ADHD), three were receiving treatment (Figure 1). Other reported mental illnesses included schizophrenia (3 cases), bipolar disorder (1 case), and personality disorder (4 cases) (**Fig. 1**).

**Figure 1:**
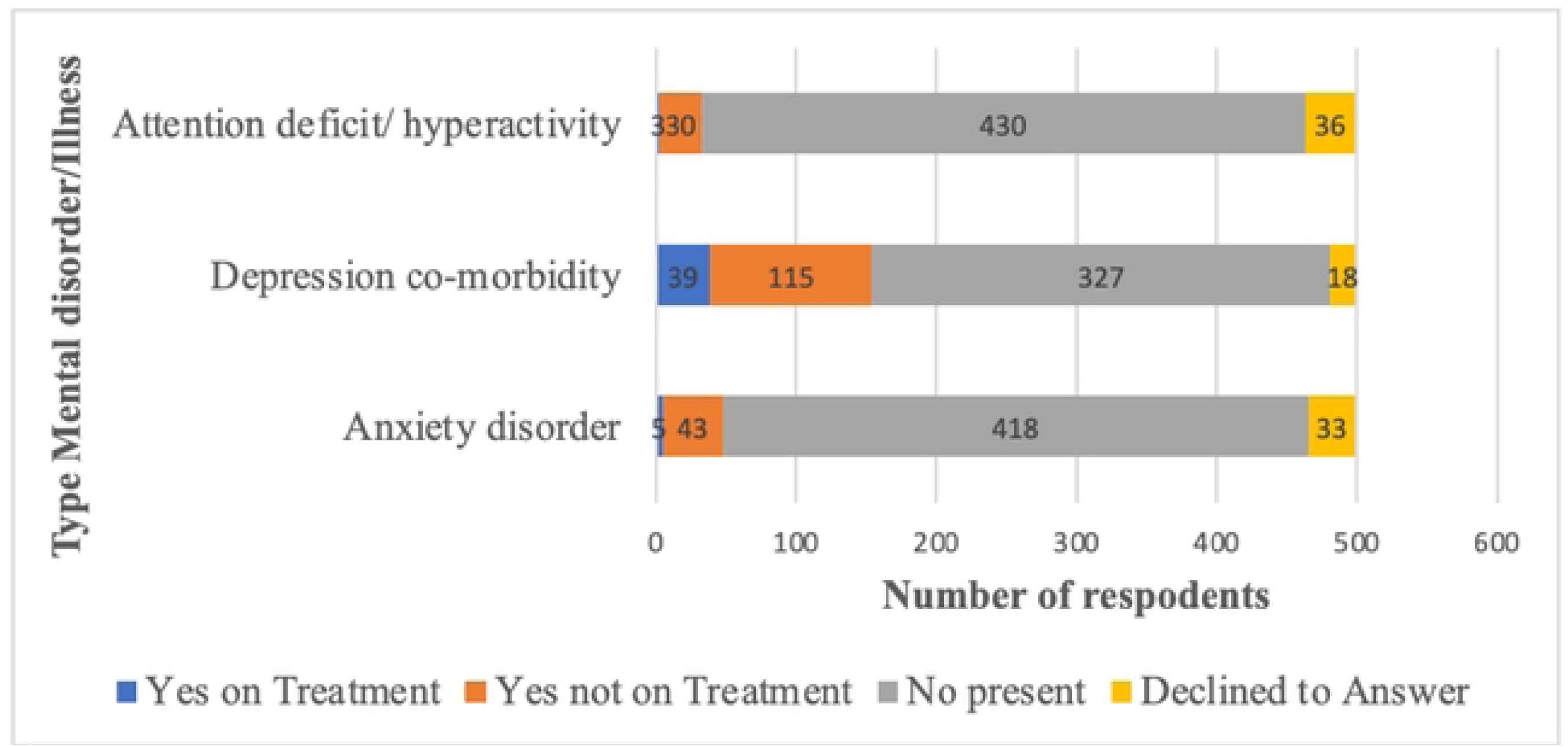
Mental Health Illnesses(self-reported) among people who use drugs in Uganda (n=499)

### Physical health illnesses

In terms of physical health, a total of 164(32.9%) participants reported experiencing fever within the three months preceding the interview, while 162(32.5%) mentioned having a cough, 21(4.2%) reported frequent urination, 34(6.8%) experienced painful urination, and 10(2%) had groin wounds. Additionally, 110 (22%) individuals had been treated for malaria, 75(15%) for sexually transmitted infections (STIs), 18 (3.6%) for urinary tract infections (UTIs), and 32(6.4%) for upper respiratory tract infections (URTIs). Five participants had received treatment for tuberculosis, and 20(4%) for skin diseases.

### Drug use-related characteristics

The median duration of drug use was .4 years (IQR: 3.9-13.8 years). Four hundred and twelve participants (82.6%), revealed that they were introduced to drug use by close friends or relatives, with 72.5% citing peer influence as the primary reason for continued drug use. Other reported reasons included coping with the loss of a loved one in 57 individuals (11.4%), coping with work-related stress(six participants), and personal illness (six) as reasons for continued drug use. Notably, 354 participants (70.9%) were categorized as PWID based on their reported history of injection drug use. Among them, 178, (50.3%) had last injected more than a month before the interview, 62 (17.5%) within the last month, 61 (17.2%) within the last week, and 21 (5.9%) on the day of the interview. Regarding injection frequency, 199 participants (57.2%) reported occasional injections, 54 (15.5%) injected at least monthly, 67 (19.3%) injected weekly, and 28 (8%) injected daily. Of the 55 participants who reported being on Medications for Opioid Use Disorder (MOUD), 35 (%) injected drugs only occasionally (**Table 2**).

**Table 2.** Drug use-related characteristics of persons who use illicit drugs in Kampala, Uganda (N = 499)

| Variable | number (%) |
| --- | --- |
| **Drug use duration median(IQR)** | **7.4(3.9-13.8)years** |
| Less than 5 years | 171 (34.3) |
| 5 to 10 years | 136(27.3) |
| Above 10 years | 192(38.5) |
| **Reasons for drug use** |  |
| Cope with the loss of a loved one | 74(14.8) |
| Cope with stress at work | 57(11.4) |
| Peer Influence | 362(72.5) |
| Cope with personal Illness | 6(1.2) |
| **Who introduced you to drug use?** |  |
| Drug dealer | 18(3.6) |
| Friend/Relative | 412(82.6) |
| Oneself | 69(13.8) |
| **Ever injected drugs** |  |
| No | 143(28.7) |
| Yes | 354(70.9) |
| Declined to answer | 2(0.4) |
| **Experience drug overdose last year** |  |
| Yes | 73(20.6) |
| No, or declined to Answer | 281(79.4) |
| **Drug injection duration median (IQR) (n=354)** | **1.8 (0.9-3.9)years** |
| **Frequency of drug injecting (n=354)** |  |
| Daily | 28(8.0) |
| Weekly | 67(19.3) |
| Monthly | 54(15.5) |
| Occasionally | 199(57.2) |
| Declined to Answer | 6(1.7) |

### HIV Risk behavior

Key HIV risk behaviors were identified, particularly related to both drug-injection and sexual risky practices in the past six months. Sharing of injecting equipment was a significant concern, with 33 participants (9.3%) admitting to using a shared needle or syringe, and a similar proportion acknowledging the sharing of other injecting materials. Additionally, 22 participants (6.2%) reported a risky practice of flashing blood, and using unsterilized syringes and needles, either on themselves or another person. Despite these risky behaviors, a majority of 268 participants (75.7%) stated that obtaining clean needles and syringes was easy, which may potentially mitigate some risks associated with injecting drug use**(Table 3)**.

**Table 3.**
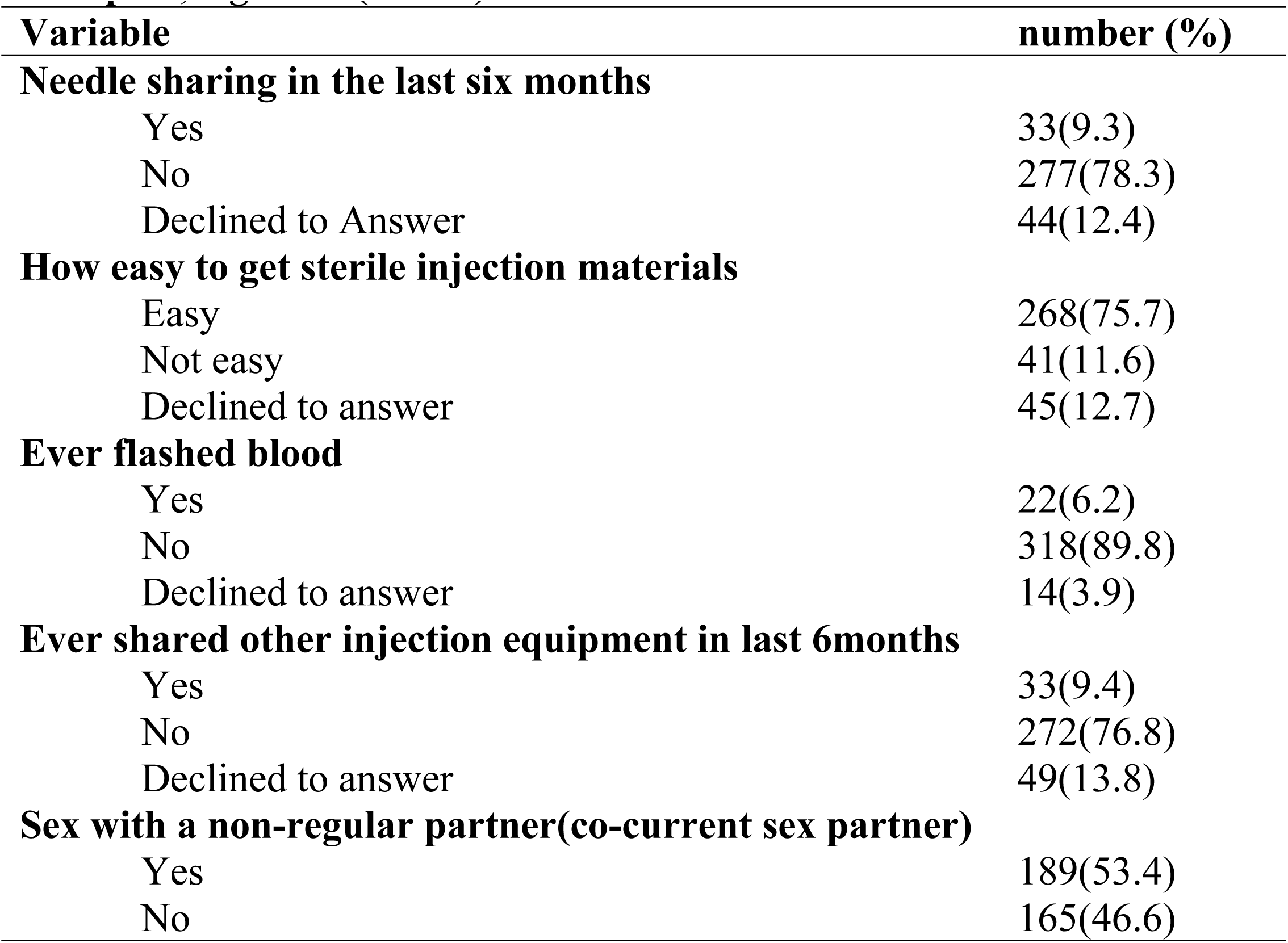
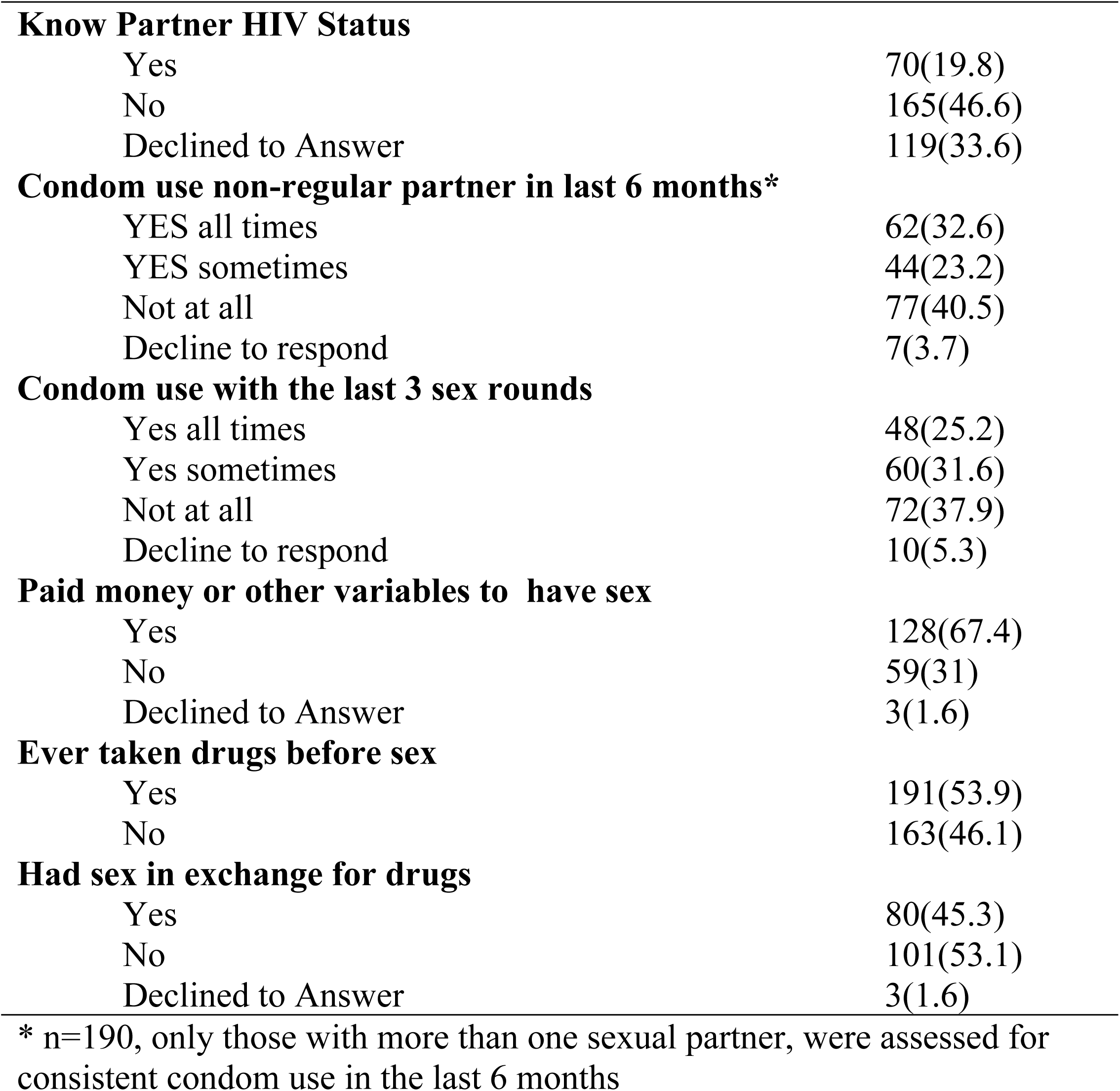
Drug use and sexual-related HIV risk factors among injection drug users in Kampala, Uganda (n=354)

### Drug use and sexual-related HIV risk factors

Regarding sexual behaviors, 189(53.4%) participants reported engaging in sexual encounters with non-regular partners within the last three months, indicating a potential pathway for HIV transmission beyond injecting drug use. Only (19.8%) knew the HIV status of their non-regular partners, suggesting a lack of awareness regarding potential HIV risks in sexual encounters. Condom use during sexual encounters with non-regular partners was low, with only 32.6% and 25.2% of participants reporting consistent condom use during sex with each non-regular partner and all their last three sexual encounters, respectively. Transactional sex was prevalent, with 128 participants (67.4%) engaging in paid or paid-for sexual encounters, and 45.3% exchanging sex for drugs. Additionally, 191(53.9%) participants reported using drugs before engaging in sexual encounters with others, indicating a heightened risk of HIV transmission due to impaired judgment and decision-making associated with drug use**(Table 3)**.

### HIV positivity among people who inject drugs

Overall, HIV status was ascertained in 354(70.9%) participants, with 13 (3.7%) self-reported to be HIV positive. HIV positivity was higher among females (8.4%) compared to males at 1.9%. A higher proportion of young persons (18-24 years) were HIV-positive (7.1%) compared to older men aged 25-34 years (2.1%) and those above 34 years (4.5%). The proportion of HIV positives was higher (16.7%) among non-Ugandans compared to Ugandan participants (3%). There was no variation in positivity based on participants’ marital status and income.

Regarding HIV risk behaviors related to drug use, HIV was more prevalent among participants who reported difficulty in accessing clean injection materials. Approximately 5.3% of those with difficult-to-access clean injection materials compared to 1% of those with ease of accessing clean injection materials reported to be HIV positive. The reported proportions of HIV positives did not vary based on duration of injection, and frequency of injecting.

In terms of sexual risk behaviors, HIV occurrence was higher among participants who reported engaging in sexual activities with non-regular partners. No variation in HIV status was reported based on other sexual risk factors such as consistent condom use, awareness of partner’s HIV status, paying money or other goods for sex, and taking drugs for sex**(Table 3)**.

### Association between socio-demographics, risk behaviors, and HIV

At bivariate analysis, female sex, non-Ugandan by nationality, difficulty in accessing sterile injection materials, and a history of taking drugs before sex, were more likely to be HIV positive. The age of the participant, having sex with a non-regular partner in the last three months, and consistent condom use with non-regular partners were not statistically significant to an individual’s HIV status (Pavlues>0.05). Multivariate analysis showed that females compared to the males were over four times more likely to report HIV-positive status {Adjusted Odds ratio(AOR)=3.8, 95% confidence interval (CI):1.01-15.5, P value=0.049}. Participants who reported having difficulty in accessing sterile injection materials were likely to be HIV positive, {AOR=4.69, 95% (CI): 1.15-19.16, p=0.031}. The other factors showed no significant association with the HIV status of the participants (**Table 4**).

**Table 4:** Association between socio-demographics, risk behaviours and HIV status of injection drug users in Kampala Uganda.

| **Variable** | **Unadjusted OR**<br>**(95% CI)** | **P-Value** | **Adjusted OR**<br>**(95% CI)** | **P-Value** |
| --- | --- | --- | --- | --- |
| **SEX** Male | 1.0 |  | 1.0 |  |
| Female | 4.67 (1.5-14.7) | 0.008 | 3.8 (0.9-15.5) | 0.049 |
| **Age** |  |  |  |  |
| 18-24yrs | 1.0 |  | 1.0 |  |
| 25-34yrs | 0.28 (0.07-1.18) | 0.082 | 0.36 (0.78-1.66) | 0.189 |
| 35yrs & above | 0.61 (0.16-2.38) | 0.48 | 1.02 (0.21-4.89) | 0.985 |
| **Nationality** |  |  |  |  |
| Ugandan | 1.0 |  |  |  |
| Non-Ugandan | 6.52 (1.62-26.18) | 0.008 | 5.24 (0.87-31.49) | 0.07 |
| **Access to sterile injection materials** |  |  |  |  |
| easy | 1.0 |  | 1.0 |  |
| not easy | 5.18 (1.56-17.18) | 0.007 | 4.69 (1.15-19.16) | 0.031 |
| decline to respond | 0.85 (0.10-7.06) | 0.878 | 0.83 (0.08-8.7) | 0.876 |
| **Sex with a non-regular partner** |  |  |  |  |
| Yes | 1.0 |  | 1.0 |  |
| No | 0.23 (0.05-1.10) | 0.067 | 0.51 (0.07-3.93) | 0.517 |
| decline to respond | 1.8 (0.21-15.48) | 0.592 | 2.6 (0.21-31.71) | 0.453 |
| **No of sexual partners in last 6 months** |  |  |  |  |
| >one Partner | 1.0 |  | 1.0 |  |
| None/one Partner | 0.61 (0.13-2.86) | 0.531 | 0.71 (0.09-5.43) | 0.746 |
| decline to respond | 0.18 (0.02-1.40) | 0.1 | 2.27 (0.17-30.03) | 0.533 |
| **Consistent condom use in last three rounds** |  |  |  |  |
| YES all times | 1.0 |  |  |  |
| YES sometimes | 0.75 (0.14-3.89) | 0.732 |  |  |
| Not at all | 0.51 (0.11-2.36) | 0.388 |  |  |
| decline to respond | 0.38 (0.74-1.95) | 0.248 |  |  |
| **Take drugs before sex** |  |  |  |  |
| Yes | 1.0 |  | 1.0 |  |
| No | 0.20 (0.04-0.93) | 0.04 | 0.51 (0.07-3.9) | 0.524 |

## Discussion

In this study, seven in ten injection drug users were males, not married, unemployed, with higher levels of education, and of varied ages. Over nine in ten were Ugandan nationals and seven in ten had ever been imprisoned. Peer influence was cited by seven in ten participants as a key determinant of drug use. Additionally, seven in ten had injected drugs in the last three months, with half reporting at least one mental health problem. One in three reported experiencing fever or cough in the last three months. Risk behaviors were common, with one in ten reporting syringe and needle sharing. Half had sex with non-regular partners, but only a third used condoms. Three-quarters engaged in transactional sex, yet only seven in ten knew their HIV status, with a positivity rate of 3.7%, higher among females and non-Ugandan nationals. Females and those with difficulty accessing sterile injection materials were more likely to be HIV positive.

Our results are comparable to an earlier descriptive studies in Uganda, that found PWID to be predominatly male, not married, with some earning a living through sex work (19). Similar findings have been reported in other regions, highlighting the vulnerability of unemployed and economically disadvantaged populations to injecting drug use (20, 21). The UNODC resource guide on counseling in targeted intervention for injecting drug users, emphasizes the importance of understanding the characterisitics of IDU, for the successful management of opioid use disorder. Further noting that IDUs are often the most vulnerable, underprivileged subgroup of the community, mainly men in their productive years, not in gainful employment due to intoxication, and frequent absenteeism at work. Reports from other developing countries similarly observed that injecting drug users were primarily men, unmarried, Christian, and unemployed(22, 23). Although the gender gap in drug use is narrowing, our findings suggest otherwise that men in Uganda and possibly similar resource-limited settings shoulder a large portion of the drug injection burden (24, 25). Peer influence was prominently cited as a predisposing factor for drug use, likely associated with the younger participants, as over 70% were below 35 years old. Young people often influence or imitate each other to demonstrate a sense of belonging. Indeed, eight out of ten participants reported that either friends or relatives introduced them to drug use. Similarly, a study among university students aged below 25 years found that nine out of ten participants identified peer influence as the main determinant of psychoactive substance use (26).

### Mental health

We found a high self-reported occurrence of mental health disorders, coupled with limited access to treatment services. Nearly half of the participants reported a diagnosis of co-existing mental disorder, with only one in ten were receiving treatment. Depression, Anxiety, and Attention Deficit Hyperactivity Disorder were the most prevalent co-morbidities. Schizophrenia, post-traumatic stress disorder (PTSD), and attention deficit hyperactivity disorder (ADHD) were less common in this population. The prevalence of mental disorders in this study was high compared to what was found in a systematic review of 24 studies from Uganda, which reported a prevalence of 24.2%, in adults (27, 28). In the same review, the combined prevalence of anxiety disorders and depressive disorders was approximately one in four adults. Globally, the prevalence of mental disorders has been found to vary widely based on the method of ascertainment, with recent studies reporting a prevalence of depressive disorders varying from as low as 3·4% to a high 66% in the general population (2). Despite potential social desirability bias associated with self-reported data, our findings suggest that the escalating injecting drug use is likely exacerbating the ongoing mental health crisis in Uganda. Diagnosing mental disorders can be challenging, particularly in low-resource settings like Uganda where the inadequate human resources to address the growing mental disorder and drug use disorder burden are limited(29). The signs and symptoms of acute intoxication and withdrawal syndrome are similar to those of other psychiatric disorders, complicating the accurate diagnosis of these conditions. Furthermore, mental disorders can be risk factors for and can exacerbate, substance use, and vice-versa, creating a complex interplay between the two(30). This interplay highlights the need for integrated approaches to address both mental health disorders and opioid use disorders in Uganda.

### Physical health profiles

Injecting drug users face a significant burden of physical health issues, including infectious diseases such as malaria, sexually transmitted infections (STIs), and HIV. The reported prevalence of malaria treatment (22%) is consistent with the high malaria burden in Uganda, as the leading cause of morbidity and mortality (31). The prevalence of STIs (15%) and HIV (4.38%) underscores the intersection of substance use and sexual health risks among this population. These findings align with other studies, which have similarly reported elevated rates of STIs, HIV and other communicable diseases, among people who use drugs. The high vulnerability has been associated with some factors including high-risk sexual behaviors associated with drug use, limitations in accessing healthcare services, and hygiene practices, among others (32, 33). The presence of tuberculosis (TB) and skin diseases among participants highlights the broader health challenges faced by PWID (25). Overall, these findings emphasize the need for integrated healthcare interventions that address both infectious and non-communicable diseases among injecting users in Uganda and similar settings.

### HIV Prevalence

The prevalence of HIV among injecting drug users was 4.38%, far less than the 33% reported by the Uganda AIDS Commission and Uganda Ministry of Health (34). The recent crane survey and two other studies found the HIV prevalence among PWID in Uganda to be ranging between 3.6% to 17% (35–37). In this study, we primarily relied on reported participants’ self-report on their HIV status, with only one in five person who had not tested in the last six months agreeing to test. It is possible that most participants who reported a negative test may have tested well before the previous three months, and some may have reported a negative HIV test due to stigma. Stigma about HIV-positive status is highly prevalent in this population, and often influences the uptake of HIV testing services(38, 39). Even with the low prevalence finding, HIV prevalence is four times higher among females than males, consistent with studies in Tanzania where the HIV-positive sero-status was three times higher in females compared to males (40). Risky injection practices and sexual behaviors contribute to the heightened risk of HIV transmission among injecting drug users(25, 41). Sharing of injecting equipment and engaging in unprotected sexual encounters with non-regular partners were common. The high frequency of transactional sex and the use of drugs before sexual encounters further compound the risk of HIV transmission. However, despite the high occurrence of risk behaviors, our study found a low HIV prevalence, likely due to the self-reported nature of HIV status. Stigma and fear may prevent persons with injecting drug use disorders from getting tested or disclosing their status. Low HIV status awareness (only one in five) among injecting drug users also suggests inadequate coverage of HIV prevention services(41). At the time of data collection, only one MOUD clinic existed in Kampala city. This highlights the urgent need to expand harm reduction services, including HIV testing, to reach more injecting drug users in Uganda.

### Risk factors for HIV infection

The study reveals the existence of risky drug injection and sexual practices in Uganda with the potential to increase the number of new HIV infections. The common risk drug injection practices reported include sharing needles and syringes, sharing mixing equipment, flushing blood, and reusing syringes multiple times. These findings reinforce earlier reports from studies in Uganda (17, 37). Prevoius studies associated highly prevalent drug risk behavior with the unaffordability of new syringes and the nadir existence of harm reduction programs in Uganda. Regarding to risky sexual practices, over half of the participants reported having sex with a nonregular partner, three of ten reported inconsistent condom use, and over two-thirds of the participants were engaging in transactional sex. These factors heighten the vulnerability of PWID to HIV infection (42). While taking drugs before sex was associated with an increased risk of HIV, this association was not significant in multivariate, similar to other sexual risk factors. A previous study by the Community Health Alliance Uganda (CHAU) examined discussed three main HIV risk perspectives: impaired judgment leading to unprotected sex, frequent sexual encounters with non-regular partners, and reduced libido due to drug use, which lowers sexual activity(37). Additionally, people who inject drugs may be forced to sell sex to acquire money to buy drugs (32, 43). In our study, nearly seven in ten of those who had multiple sexual encounters, paid money or other commodities in exchange of sex with another person, and nearly half reported having had sex in exchange for drugs.

Another crucial risk factor deserving attention is the status of being a non-Ugandan, primarily refugees, who may have recently imigrated. The risk of being HIV positive was found to be five times higher among non-nationals compared to Ugandans. This aligns with findings from studies conducted in Dar-es-Salaam and the United States, where recent immigrants exhibited an increased risk of HIV infection (6, 44). We attribute this heightened risk among non-nationals to various factors, including unawareness of safer needle use practices, financial constraints hindering the purchase of new needles, and exposure to circumstances that increase the likelihood of engaging in risky sex work. The small sample size and missing data on key parameters may partially explain the failure to demonstrate an association between injection drug risk and the HIV status. Despite efforts to ensure the confidentiality and safety of participants during interviews, it was not uncommon for participants to decline to respond to particular essential questions or take an HIV test. Another factor that we didn’t find significant was the frequency of injection; in Uganda, most drug users only injected occasionally. This affects the risk of exposure to HIV and programming for harm reduction interventions such as clean needle and syringe distribution. Nonetheless, our study finding reinforces current evidence on the relation between HIV serostatus, and injecting drug and sexual risk practices. Two studies conducted in Uganda found that participants reused syringes in some instances up to 2-4 times before disposal (17, 45).

### Limitations

Selection bias may have emerged due to increased security enforcement in preparation for a Non-Aligned Movement (NAM) Summit meeting held in Kampala, during January 2024, coinciding with our data collection period. This security exercise drove more PWID into hiding, reducing our access to some community members. Selecting initial seeds on a first-come basis likely favored accessible or motivated individuals, leading to an unrepresentative sample. This may underrepresent those more stigmatized, severely addicted, or with poor health and less access to services, skewing results and underestimating the true prevalence of risk behaviors and mental and physical health conditions.

In some instances, participants declined to answer or did not respond to a question. Although this could have been because individuals deliberately preferred not to answer, we can’t rule out the fact that cognitive impairments, mood disorders, or other mental health issues suffered by people suffering from opioid use disorder, could compromise the ability to recall and report information accurately.

Relying on self-reporting by participants could have introduced recall bias, as participants might not accurately remember past events or might selectively report on such events, leading to misrepresentation of the true measures. To minimize this bias, the questions were designed with a maximum recall duration of three months, pretested, and simplified to reduce cognitive load and facilitate easier recall of past events. Additionally, interviews were rescheduled based on participants’ availability to enhance recall accuracy, avoid distractions, and allow for better concentration and memory recall. Furthermore, while measures were taken to ensure confidentiality and safety, the possibility of overreporting or underreporting drug and sexual risk behaviors remains, as participants may have been influenced by social desirability bias stemming from internal stigma. The extended data collection period(six months) allowed us to reach and enroll injecting drug users of different categories. Collecting data at 38 community hotspots where most injecting drug users congregate helped ensure diversity. Using ODK enabled real-time data correction, increasing accuracy. Based on these adjustments, we believe these results are valid and representative of injecting drug users in Kampala.

### Conclusion

Our study provides a comprehensive insight into the socio-demographic, mental, physical health, and HIV-associated risk of persons with drug user disorders in Kampala, Uganda. The findings indicate a significant vulnerability to injecting drug use, mental health disorders, and high-risk behaviors that predispose this population to HIV infection. Despite a low HIV prevalence compared to previous estimates, the interplay between drug use, risky injecting practices, and sexual behaviors suggests an urgent need for targeted interventions to address these intertwined challenges. Implementing a multi-sectoral approach that combines, harm reduction, physical healthcare, and mental health services is crucial for improving the well-being of persons who inject illicit drugs, and reducing the burden of HIV and other associated health issues in Uganda.

## Data Availability

The datasets analysed during the current study will be shared on reasonable request by the corresponding

## Supporting information Abbreviations

CHAU: Community Health Alliance Uganda
MOUD: medication for opioid use disorder
OUD: Opioid use disorder
PWID: People Who inject drugs
IDU: Injecting drug user
HRN: Uganda Harm Reduction Network Organization

## Acknowledgment

The authors gratefully acknowledge the support from the Butabika National Referral Mental Hospital management and MOUD clinic staff who participated in the data collection and cleaning. Special thanks go to the Ugandan government and the United States PEPFAR Uganda Centers for Disease Control and Prevention (CDC) for supporting the establishment of the MOUD program in the country.

